# Examining the Accuracy and Reproducibility of Responses to Nutrition Questions Related to Inflammatory Bowel Disease by Generative Pre-trained Transformer-4 (GPT-4)

**DOI:** 10.1101/2023.10.28.23297723

**Authors:** Jamil S. Samaan, Kelly Issokson, Erin Feldman, Christina Fasulo, Nithya Rajeev, Wee Han Ng, Barbara Hollander, Yee Hui Yeo, Eric Vasiliauskas

## Abstract

**Background and Aims:** Generative Pre-trained Transformer-4 (GPT-4) is a large language model (LLM) trained on a vast corpus of data, including the medical literature. Nutrition plays an important role in managing inflammatory bowel disease (IBD), with an unmet need for nutrition-related patient education resources. This study examines the accuracy, comprehensiveness, and reproducibility of responses by GPT-4 to patient nutrition questions related to IBD.

**Methods:** Questions were obtained from adult IBD clinic visits, Facebook, and Reddit. Two IBD-focused registered dieticians independently graded the accuracy and reproducibility of GPT-4’s responses while a third senior IBD-focused registered dietitian arbitrated. Each question was inputted twice into the model.

**Results:** 88 questions were selected. The model correctly responded to 73/88 questions (83.0%), with 61 (69.0%) graded as comprehensive. 15/88 (17%) responses were graded as mixed with correct and incorrect/outdated data. The model comprehensively responded to 10 (62.5%) questions related to “Nutrition and diet needs for surgery”, 12 (92.3%) “Tube feeding and parenteral nutrition”, 11 (64.7%) “General diet questions”, 10 (50%) “Diet for reducing symptoms/inflammation” and 18 (81.8%) “Micronutrients/supplementation needs”. The model provided reproducible responses to 81/88 (92.0%) questions.

**Conclusion:** GPT-4 comprehensively answered most questions, demonstrating the promising potential of LLMs as supplementary tools for IBD patients seeking nutrition-related information. However, 17% of responses contained incorrect information, highlighting the need for continuous refinement prior to incorporation into clinical practice. Future studies should emphasize leveraging LLMs to enhance patient outcomes and promoting patient and healthcare professional proficiency in using LLMs to maximize their efficacy.

**Lay Summary:** Generative Pre-trained Transformer-4 (GPT-4) is a large language model that comprehensively answered patient nutrition questions related to IBD. With continuous refinement and validation, there is promising potential for GPT-4 in enhancing outcomes and promoting health literacy in this patient population.

## INTRODUCTION

Inflammatory bowel disease (IBD), which includes Crohn’s disease (CD) and ulcerative colitis (UC), are chronic, often debilitating conditions impacting an estimated 4.9 million cases worldwide (1). Prevalence is highest in the United States and China with studies showing a rising incidence in newly industrialized nations that have adopted more Western lifestyles (1,2) Nutrition plays an important role in the management of patients with IBD, not only influencing disease activity but also impacting overall patient outcomes given the increased risk for malnutrition and micronutrient deficiencies in this patient population.(3,4) Proper nutritional interventions, such as exclusive enteral nutrition (EEN) can alleviate symptoms, reduce the need for medications, and promote mucosal healing, thus improving their quality of life.(4–7) Furthermore, diet and lifestyle modifications can influence the course of these diseases by modulating gut microbiota, inflammatory responses, and disease-related complications. (8)

The important role of nutrition in IBD underscores a pressing need for effective patient education in this domain. Patient surveys have consistently indicated a strong demand for comprehensive nutrition guidance and education, both in general and within the IBD community.(9) One study of IBD patients showed 15.4% (86/559) of patients did not talk to any provider about nutrition while 15.5% (63/407) rarely entered discussion with their healthcare providers regarding this subject. When asked the reason for the lack of discussion, 31.3% (21/67) of patients felt their provider did not have enough time during clinical visits while 20.9% (14/67) reported feeling their provider lacked the requisite expertise on the subject.(10) Moreover, studies show patients frequently turn to external sources of information in lieu of their healthcare providers.(11,12) More and more patients are seeking information about their disease from online sources, and this trend is anticipated to grow.(13) Despite this demand, there is variable access to high quality IBD-specific dietary information available to patients.(14) Furthermore, when navigating online sources patients are confronted with both high-quality information as well as misinformation that may be harmful, a dichotomy that often makes it challenging to distinguish between valid medical guidance from erroneous or misleading advice.

Large language models (LLMs), like ChatGPT, may revolutionize the way information is accessed and disseminated. These models have the potential in assisting patients with IBD by helping them navigate the complex landscape of nutrition information. Personalized dietary recommendations can play a pivotal role in managing IBD, and LLMs may offer an accessible platform for patients to receive timely and pertinent information. While not a replacement for professional medical advice, LLMs can serve as a complementary resource by providing general dietary guidelines and addressing common nutritional queries. Studies have shown the impressive ability of ChatGPT in answering clinical questions on a wide range of topics including cirrhosis, bariatric surgery, laryngology and orthopedic surgery. (15–18) Studies have also examined the performance of LLMs in gastroenterology with a recent study showing GPT-4 surpassed the passing grade threshold and average human test taker performance on the American College of Gastroenterology self-assessment.(19) There is a growing body of evidence demonstrating the strength and limitations of LLMs across multiple topics in gastroenterology including IBD.(20–24) One recent study showed ChatGPT has promise in providing appropriate answers to questions related to the dietary management of IBD although the study was limited by a small number of questions evaluated (n=6). (25)

This study aims to expand on the current literature by examining the accuracy, comprehensiveness, and reproducibility of ChatGPT when answering nutrition-related questions in IBD. Examining the performance of LLMs, like ChatGPT, in the realm of nutrition will be important to understanding the future role and limitations of this technology.

## MATERIALS AND METHODS

### Question Selection/Data Source

All questions posed during adult patient clinic visits with IBD focused registered dietitians at a tertiary academic hospital, Facebook groups (“Crohn’s and Colitis Food, recipes, Diet, and Nutrition”, “Crohn’s Disease & Ulcerative Colitis Support Group”, “Crohn’s Disease Support Group”) and Reddit pages (“r/CrohnsDisease”, “r/IBD”) during the month of February 2023 were considered for inclusion. Questions were obtained, screened, and approved by three authors (CF, EF, KI) to evaluate their inclusion in the study. Only nutrition questions related to IBD were selected. Duplicate and similar questions from multiple sources were removed. Questions requiring subjective or personalized responses (ex.) and questions that were vague (ex.) were rephrased to a generic language format to allow for inclusion in the study. Other questions were grammatically modified to eliminate ambiguity. A total of 88 questions were selected and used to generate responses from GPT-4. To better characterize GPT-4’s performance in various topics within IBD nutrition, questions were categorized into multiple groups for statistical analysis purposes: 1. Nutrition and diet needs for surgery, 2. Tube feeding and parenteral nutrition 3. General diet questions, 4. Diet for reducing symptoms/inflammation and 5. Micronutrients/Supplementation needs. Approval from the institutional review board was not sought given all questions are publicly available.

### ChatGPT

ChatGPT is an LLM trained using an expansive dataset from various sources such as online websites, books, and articles up to the year 2021 at the time of data collection, including the medical literature.(26) GPT-4, or Generative Pre-trained Transformer 4, was released in March of 2023 and is the fourth iteration of the GPT series. Based on a given input, the model generates responses that are easy-to-understand and conversational in nature.(27) By leveraging billions of sentences from diverse internet sources, GPT-4 can engage in complex conversations, answer questions, write essays, and even generate creative content like stories or poems. Its developers employed Reinforcement Learning from Human Feedback (RLHF) to optimize the model, ensuring it effectively responds to a wide range of commands and written inquiries. This fine-tuning was driven by human preferences, which served as a reward signal, ensuring the model’s alignment with user intent and desired outcomes.(28) The model has been fine-tuned to resonate with user goals and to reduce biased, or potentially harmful outputs. The exact origins of the data used to train ChatGPT remain undisclosed. As a product of extensive research and refinement, this model has found applications across various industries with growing literature highlighting its potential use in the field of healthcare.

### Response Generation

Each question was prompted to the March 14^th^ 2023 version of GPT-4 to generate responses. GPT-4 was accessed using the OpenAI website interface. Default model settings (temperature, max token size, etc.) were used. Questions were entered into the model without additional prompting in order to emulate the manner in which patients may potentially engage with the model. Every question was entered twice at different times using the “new chat” feature, yielding two responses for each question. This approach aimed to assess the reproducibility of responses to the same question.

### Question Grading

Responses to the questions underwent an initial evaluation for accuracy, comprehensiveness, and reproducibility by two actively practicing IBD-focused registered dietitians in a tertiary care center (E.F., C.F.). In instances of disagreement regarding reproducibility or accuracy grading, a third reviewer who has 10 years of experience as a clinical dietitian working with IBD patients at a tertiary care center arbitrated (K.I.). Given GPT-4’s training data was limited to information prior to September 2021, the reviewers used this date as a benchmark for assessing accuracy and comprehensiveness. This meant, the model was not critiqued on any guidelines (American, European, etc.) or scientific literature published after this date.

Accuracy and comprehensiveness were graded using a grading scale used in previous studies(15,16,29–31):

- Comprehensive: Defined as a response meeting two criteria: 1) contains only accurate information without any inaccurate information, 2) contains comprehensive information, nothing more a registered dietitian would add if asked this question by a patient in clinic.
- Correct but inadequate: Defined as a response meeting two criteria: 1) contains only accurate information without any inaccurate information, 2) does not contain comprehensive information, a registered dietitian would have more important information to add if asked this question by a patient in clinic.
- Mixed with correct and incorrect/outdated data
- Completely incorrect: No correct information was provided.

Reproducibility was assessed by examining the similarity in accuracy between the two responses provided for each question. To assess reproducibility, both responses for each question were categorized into two categories based on the presence of incorrect information: grades comprehensive and correct but inadequate formed the first category, while grades of mixed with correct and incorrect/outdated data and completely incorrect made up the second. A pair of responses to a question were deemed non-reproducible if their respective grades belonged to separate categories.

### Statistical Analysis

The proportions of responses earning each grade were calculated and shown as counts and percentages. All analyses were conducted using Microsoft Excel. (version 16.69.1).

## RESULTS

In total, 88 nutrition questions related to IBD were inputted into GPT-4. The model performed well overall in terms of accuracy and provided correct responses to 73/88 questions (83.0%), with 61 (69.0%) of responses graded as comprehensive. For example, the model provided a comprehensive summary of EEN that is clear, concise, comprehensive, and accurate. This included a summary of the route of administration, avoidance of consumption of solids, nutritional content of EEN, its impact on mucosal healing, length of administration, reintroduction of solids, and its efficacy in inducing remission particularly in children and adolescents. Another example of a comprehensive response is an important and common questions asked by patients regarding taking vitamins and minerals despite normal laboratory values. The model first stressed the importance of monitoring nutrient deficiencies in patients with IBD and the temptation by patients to discontinue these supplements when laboratory tests reveal normal values. The model then advised the user to consult with their healthcare professional who will consider the patient’s medical history, medications, and IBD disease status prior to making such a decision.

A total of 15/88 (17.0%) responses were graded as mixed with correct and incorrect/outdated data while no responses were graded as completely incorrect. For example, the model was asked about diet recommendations for IBD patients, which is one of the most common questions encountered in the IBD clinic. The model offered various suggestions that are not necessarily applicable to all patients with inflammatory bowel disease. For example, a low-fat and lactose-free diet may only be suitable for a limited subset of these patients. Furthermore, the model recommended a low residue diet, often conflated with “low fiber” diet, which is not recognized as a diet by the Academy of Nutrition and Dietetics due to the lack of definition and therefore inability to accurately measure from available data(32). This also contradicts the current International Organization for the Study of IBD (IOIBD) Diet Guidelines, which recommend patients with IBD consume more fiber. These recommendations may lead to confusion or unnecessary nutritional restrictions for patients and ultimately to worse outcomes. Another example is the model stating “a plant-based diet is not a guaranteed cure” when asked if a plant-based diet is a cure for IBD. This is inaccurate as it suggests that a plant-based diet may sometimes cure IBD.

When examined by question category, the model provided comprehensive responses to 10 (62.5%) questions related to “Nutrition and diet needs for surgery”, 12 (92.3%) “Tube feeding and parenteral nutrition”, 11 (64.7%) “General diet questions”, (50%) “Diet for reducing symptoms/inflammation” and 18 (81.8%) to “Micronutrients/Supplementation needs” (Table 1, Figure 1). The percentage of comprehensive responses was lowest among questions regarding “Diet for reducing symptoms/inflammation” at 50% and highest among questions regarding “Tube feeding and parenteral nutrition” at 92.3%.

**Figure 1:**
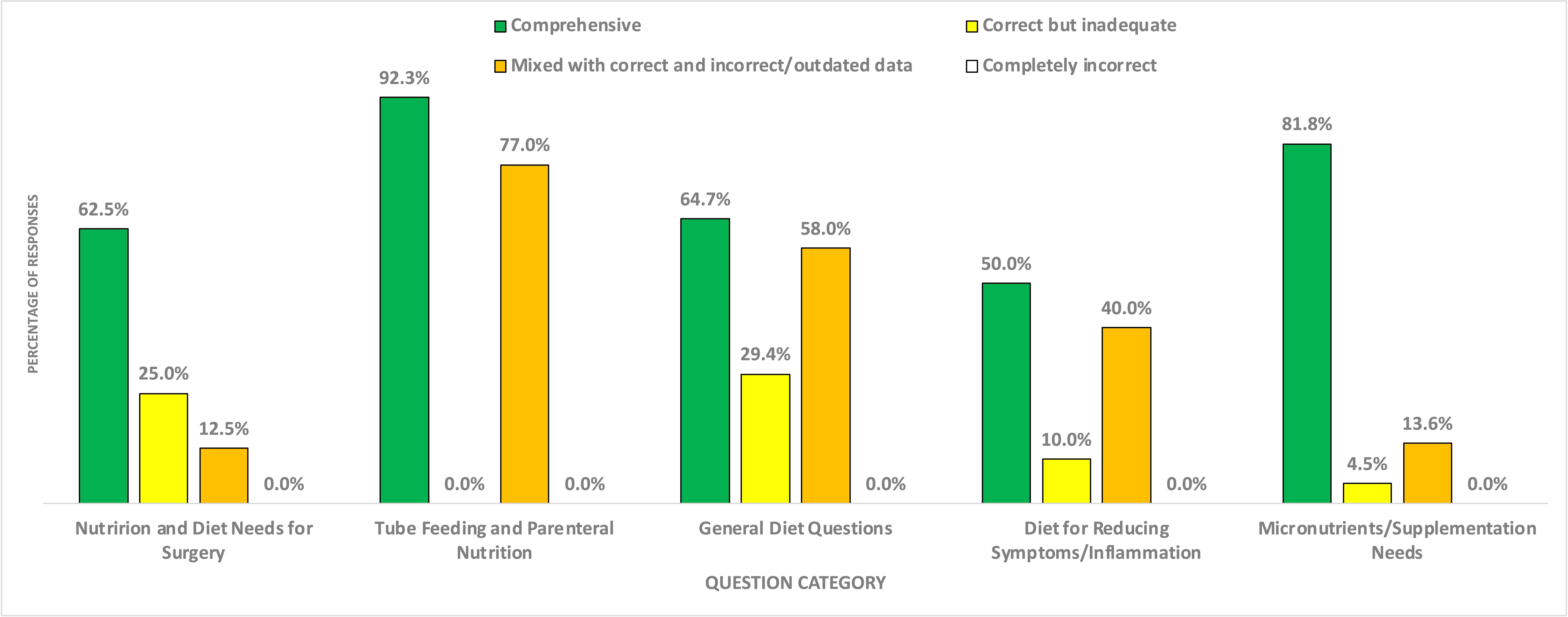
Graphical illustration of grading of responses generated by GPT-4 to nutrition questions related to inflammatory bowel disease stratified by question category.

**Table 1:**
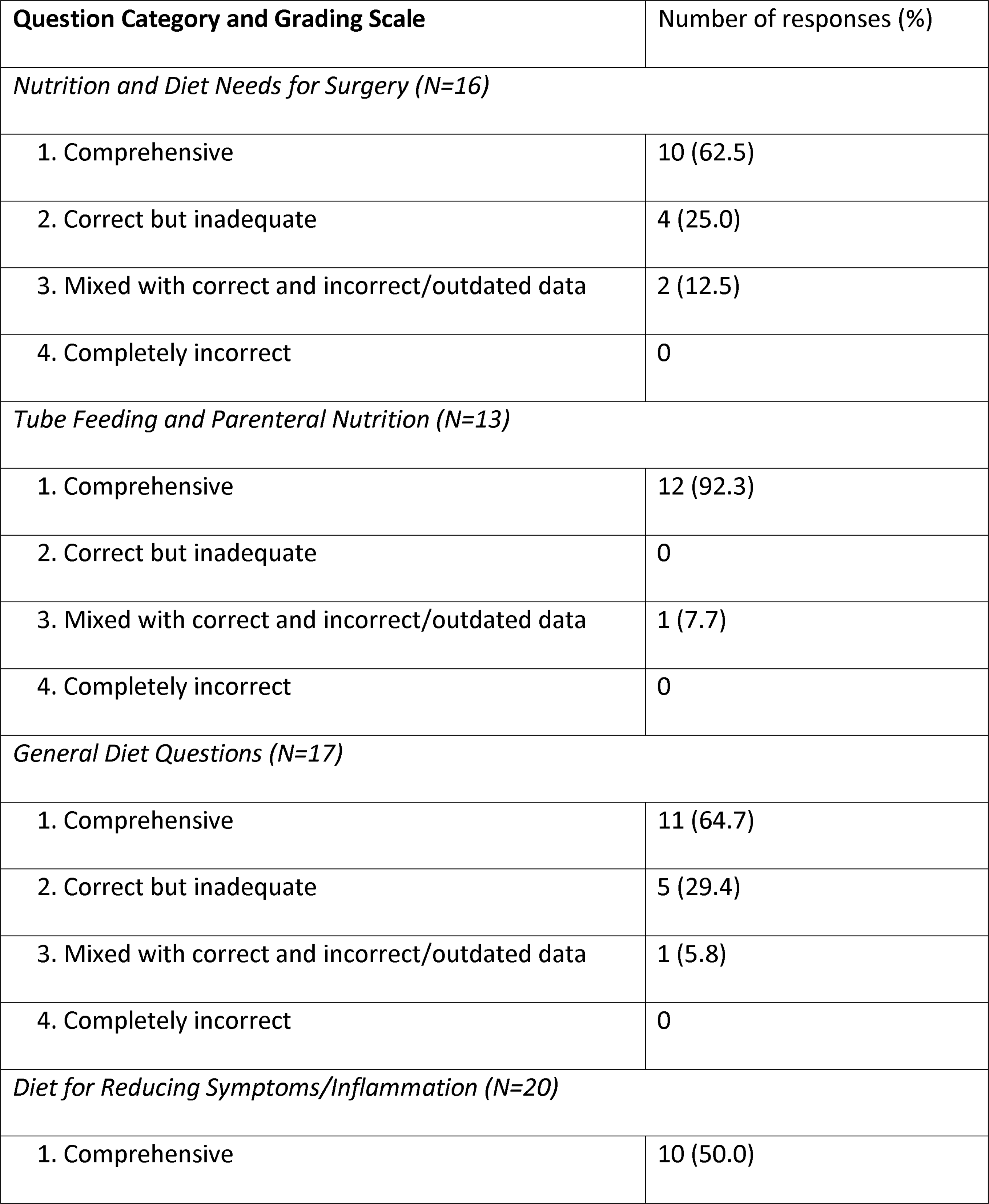
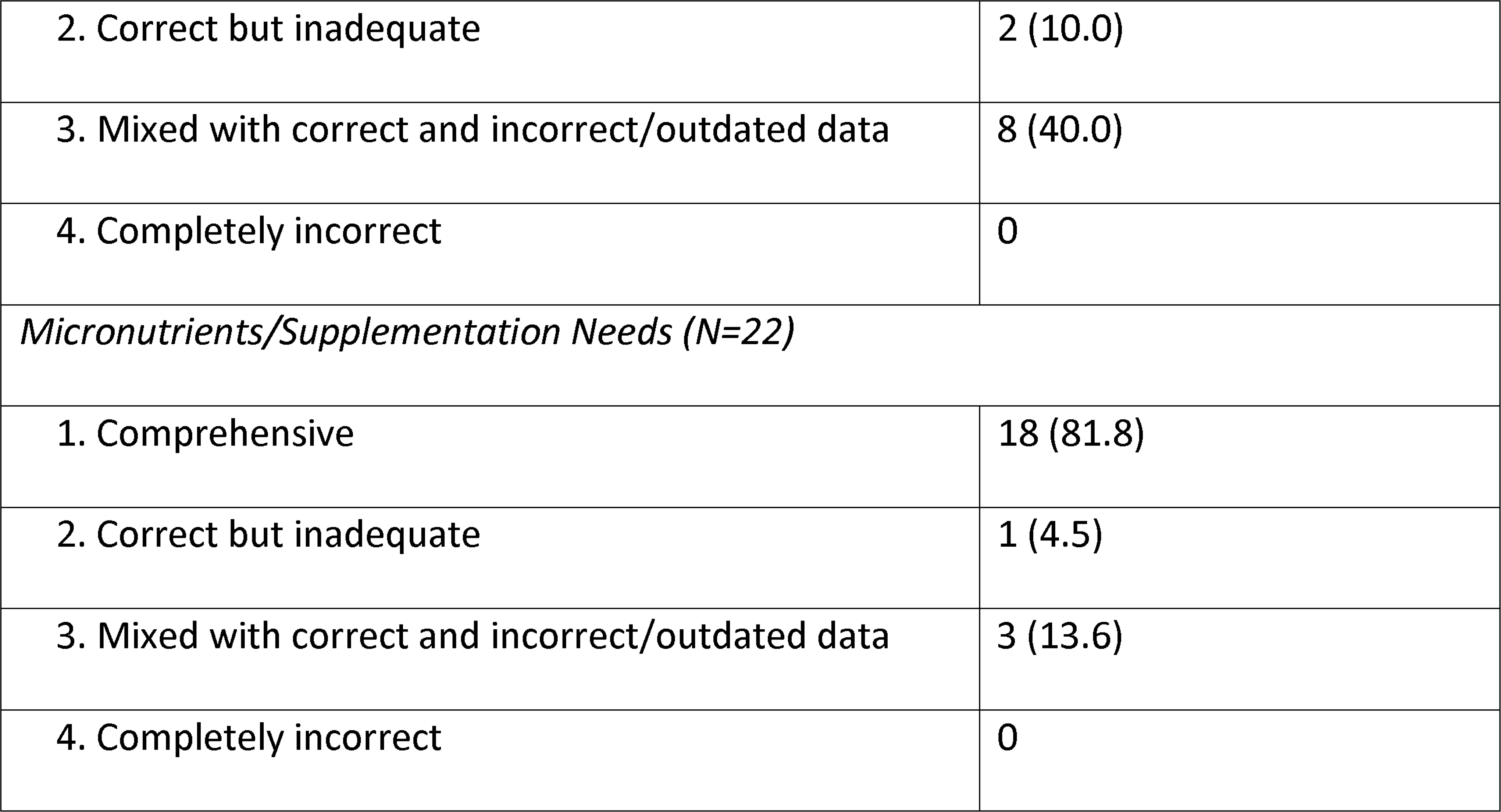
Grading of responses generated by GPT-4 to nutrition questions related to inflammatory bowel disease stratified by question category.

The model provided overall high reproducibility in accuracy with 81/88 (92.0%) of questions generating reproducible responses. When examined by question category, the model provided reproducible responses to 100% of questions related to “Tube feeding and parenteral nutrition” (100%), 94.1% “General diet questions”, 90.0% “Diet for reducing symptoms/inflammation”, 95.4% “Micronutrients/Supplementation needs”, and 81.3% “Nutrition and diet needs for surgery” (Table 2).

**Table 2.**
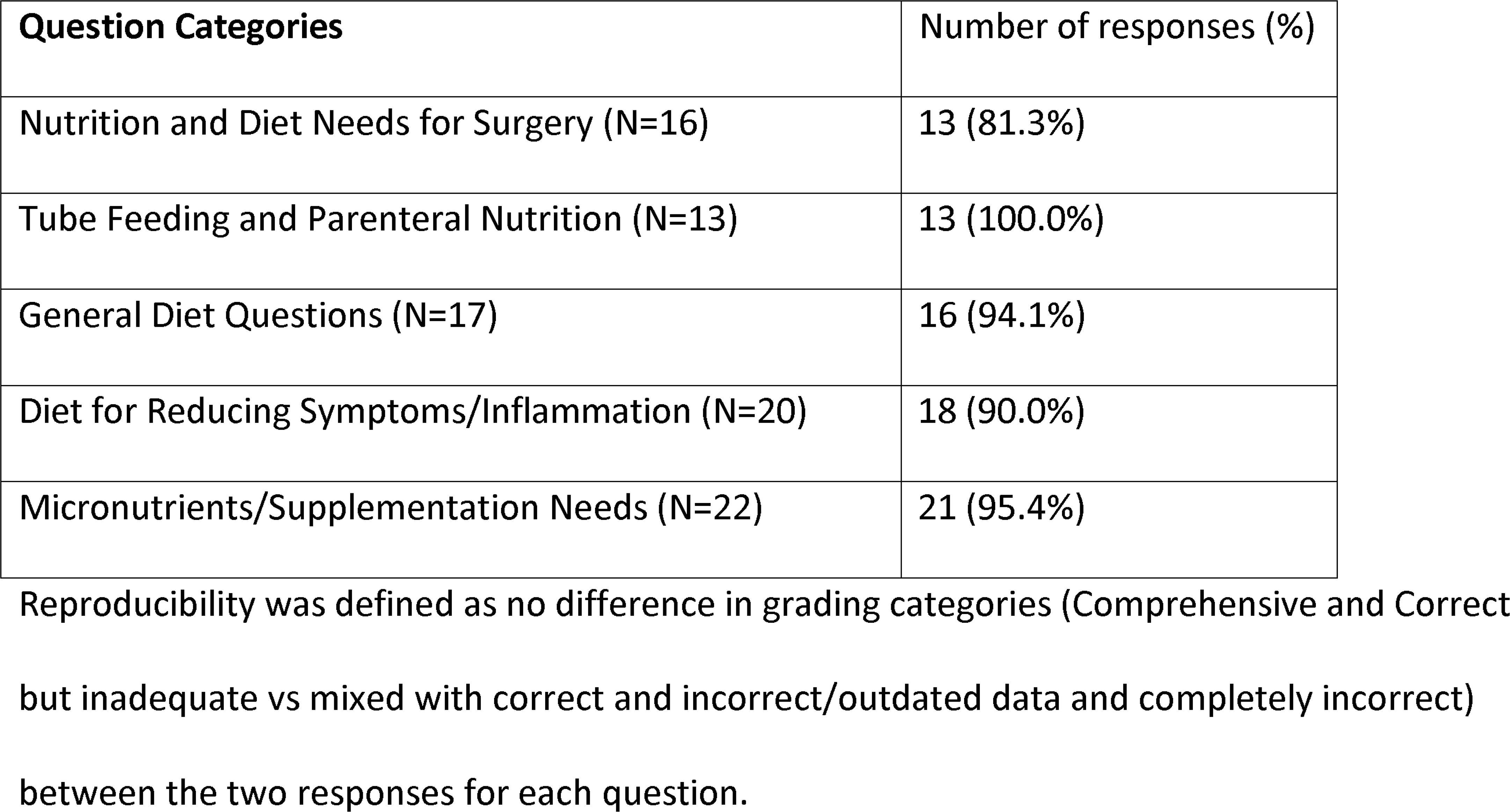
Proportion of nutrition questions related to inflammatory bowel disease with reproducible grading of responses generated by GPT-4 stratified by question category.

## DISCUSSION

The increasing global prevalence of IBD, coupled with the important role of nutrition in its management, underscores the importance of reliable and accessible nutrition-related patient educational resources. Our study evaluated the accuracy, comprehensiveness, and reproducibility of GPT-4’s responses to patient nutrition questions related to IBD. The model provided accurate and reproducible responses to the majority of questions prompted, highlighting the future potential of LLMs as a complementary source of information related to nutrition for IBD patients. However, the model provided inaccurate information in 17% of responses, underscoring the need for further improvement in performance prior to its introduction into clinical practice. With future iterations and improvements in performance, we see this technology as a supplementary tool rather than a substitute for advice from licensed healthcare professionals.

Individuals with IBD place significant emphasis on their diet, frequently implementing self-imposed dietary limitations(33–35). Subsequently, patients frequently seek nutritional guidance and turn to external sources in lieu of their healthcare providers with one study revealing that IBD patients were most confident in advice from online sources when experiencing active disease(11,12,33). The quality and reliability of information available online are either limited or frequently inconsistent. Using conventional online search engines provides patients with an abundance of information from a variety of sources, making distinguishing reliable information both difficult and time consuming. As the disease predominantly affects young adults with high computer literacy, the potential impact of online resources, like LLMs, becomes increasingly significant. This provides urgency in examining the role of LLMs in nutrition related patient education in order to understand the strengths and limitations of this technology. A recent study evaluated the accuracy of three LLMs, including ChatGPT, in answering 6 IBD dietary management questions with promising results. Two physician reviewers found ChatGPT answered most questions appropriately.(25) Our study builds on these findings with a larger sample of questions as well as utilizing actively practicing academic dietitian reviewers. GPT-4 provided accurate and comprehensive responses to the majority of questions in our study, highlighting a promising potential for LLMs in the realm of patient education. The reproducibility of GPT-4’s responses in our study, which exceeded 80% across all categories, is also noteworthy and crucial for building trust among users and ensuring consistently accurate information. If validated, LLMs like ChatGPT may serve as an easy-to-understand and efficient tool for patients to obtain reliable supplemental information based on the medical literature.

Its also important to highlight the limitations of GPT-4, as seen in our study. The model provided incorrect or outdated information in 17% of responses which demonstrates its lack of readiness for clinical use in its current form. There are multiple possible reasons for this. ChatGPT’s training includes data from a wide range of sources, some of which may not be accurate, leading to potential inaccuracies in its outputs. Moreover, the data sources used to train the model have not been publicly disclosed making critical appraisal of its knowledge base regarding a particular topic not possible. Even when accessing information from the medical literature in its training data, the model may emphasize or utilize conflicting or outdated information resulting in inaccurate or inconsistent outputs. Prompting strategies may also play a role in the quality of responses. For our investigation, we pursued a pragmatic study design where real world patient questions were obtained from our IBD clinic and online sources and subsequently inputted into GPT-4 without additional prompting. This is likely not the optimal prompting method to produce the highest quality outputs from LLMs. We hypothesize that more advanced prompt engineering would likely produce higher quality responses, both in accuracy and comprehensiveness. In light of this, the design of intuitive, precise prompts for both patients and healthcare providers may be an effective strategy in harnessing the full potential of LLMs, as it ensures these tools deliver relevant, accurate information to specific medical contexts. By educating both groups on effective prompt crafting, we can greatly enhance the quality of interactions with LLMs, potentially leading to improved patient outcomes, more efficient healthcare delivery, and a deeper integration of AI in healthcare.

If future iterations of this technology are validated for clinical use, they may have a significant impact on the patient experience and healthcare outcomes. The timely accessibility of healthcare providers, particularly dietitians and IBD specialists, remains a consistent hurdle for many IBD patients.(36–38) This can not only exacerbate delays in care and result in potentially detrimental outcomes, but also augment the challenges of patients navigating through the sea of inaccurate or insufficient online information. These barriers may also disrupt the proactive efforts of informed patients, leading to misguided actions based on flawed knowledge. In the future, patients equipped with accurate and holistic knowledge from LLMs may be able to streamline in-person appointments with more focused questions, allowing healthcare providers to focus on patient-specific concerns.(39) Moreover, empowering patients with more information can potentially foster a greater sense of autonomy among patients, encouraging them to initiate discussions regarding nutrition with their healthcare providers. This, in turn, may help promote the focus on nutrition during clinic visits, and potentially increase the rate of referrals to dietitians.

LLMs may also help bridge healthcare disparities by democratizing access to health information. Barriers to care, such as lengthy wait times and inaccessibility to a dietician or IBD specialist, are even more pronounced in patients of minority backgrounds, which can further exaggerate disparities and lead to delays in diagnosis and treatment.(40) LLMs may empower these patients with knowledge regarding their disease, giving them a tool which helps them proactively engage with their healthcare providers. LLMs may also prove beneficial to patients who have language discordance with their healthcare providers, given disparities in healthcare outcomes based on language preference have been previously shown.(41,42) ChatGPT’s ability to provide patient education in languages other than English is an active area of research with some studies demonstrating its ability to respond to inquiries related to cirrhosis in Arabic, Korean, Mandarin, and Spanish (31,43,44). LLMs can provide a valuable resource for these patients, and potentially serve to mitigate disparities in outcomes.

### Limitations and Future Directions

This study had several limitations. First, the selection of 88 questions, though reflective of common patient inquiries, was not exhaustive and did not encompass all possible topics that patients may ask. Modifications to the grammar or context of certain questions were made to improve the clarity for GPT-4, which does not reflect the variability and ambiguity often present in patient inquiries in real-world settings. In practice, LLMs have the ability to request clarification from users, a feature that was not incorporated into the study design, potentially affecting the model’s performance under real-world conditions. Duplicate questions were removed during screening which introduced the potential for selection bias. LLMs may respond differently to different wording of the same question. Another limitation is the subjective nature of the grading scale and reviewers used in our study. Although experienced registered dietitians independently graded the responses, future research should prioritize the development of more objective and standardized methods to evaluate accuracy, comprehensiveness, and reproducibility. Moreover, only registered dietitians were involved in the review process, which may limit the scope of the evaluation. Future studies would benefit from a multidisciplinary panel of healthcare provider reviewers in addition to patients, to provide a more holistic assessment of the quality and relevance of GPT-4’s responses.

Looking ahead, further avenues of research should aim towards ameliorating the inaccuracies and limitations of this technology as revealed by our study. While ChatGPT can provide answers to general IBD nutrition-related questions, it lacks the ability to comprehensively assess individual health status, address behavioral needs, or monitor progress over time. IBD can vary significantly from person to person in terms of symptoms, triggers, and nutritional needs. Registered dietitians can assess individual cases, considering medical history, current medications, symptom severity, and tailor nutrition advice to the individuals’ specific needs and goals. While the model’s linguistic prowess is commendable, it can sometimes produce responses that sound convincing but may be incorrect or nonsensical, a phenomenon termed “hallucinations”. Continuous feedback and iterative improvements are essential to minimize these limitations. Considering the growing popularity of LLMs and their potential in delivering patient education, it’s imperative to further study and validate their utility. The role of dieticians and physicians in managing patients with IBD remains critical to positive outcomes, and further research should investigate ways in which collaborations between AI-models and the healthcare team can benefit our patients.

## CONCLUSION

GPT-4 provided accurate and comprehensive responses to the majority of nutrition questions related to IBD, demonstrating the promising potential of LLMs as supplementary tools for IBD patients seeking nutrition-related information. It’s important to note that 17% of responses contained incorrect information, highlighting the need for continuous refinement and validation of LLMs prior to their incorporation into clinical practice. Going forward, it’s essential to approach the use of LLMs as an adjunct to professional medical advice. Future studies should focus on leveraging LLMs to enhance patient outcomes in the realm of IBD nutrition. Furthermore, efforts towards promoting patient and healthcare professional proficiency in using LLMs are essential to maximizing their impact and personalization.

## Ethical Approval

This article does not contain any studies with human participants or animals performed by any of the authors.

## Informed Consent Statement

Informed Consent does not apply.

## Declaration of AI and AI-assisted technologies in the writing process

During the preparation of this work the authors used GPT-4 in order to improve readability and language. After using this tool/service, the authors reviewed and edited the content as needed and take full responsibility for the content of the publication.

## Statement of Data Availability

Data not publicly available.

## Author contributions

J.S.S conceived and designed the study, manuscript preparation.

K.I. Question curation, response grading, edited the paper for important intellectual content.

E.F. Question curation, response grading, edited the paper for important intellectual content.

C.F. Question curation, response grading, edited the paper for important intellectual content.

W.N. Data management, statistical analysis, edited the paper for important intellectual content.

N.R. Assisted with manuscript preparation, edited the paper for important intellectual content.

B.H. Edited the paper for important intellectual content.

Y.Y. Edited the paper for important intellectual content.

E.V. Edited the paper for important intellectual content.

## Data Availability

All data produced in the present study are available upon reasonable request to the authors

## REFERENCES

1. Wang R, Li Z, Liu S, Zhang D. Global, regional and national burden of inflammatory bowel disease in 204 countries and territories from 1990 to 2019: a systematic analysis based on the Global Burden of Disease Study 2019. BMJ Open. 2023 Mar;13(3):e065186.

2. Ng SC, Shi HY, Hamidi N, Underwood FE, Tang W, Benchimol EI, et al. Worldwide incidence and prevalence of inflammatory bowel disease in the 21st century: a systematic review of population-based studies. The Lancet. 2017 Dec;390(10114):2769–78.

3. Hashash JG, Elkins J, Lewis JD, Binion DG. AGA Clinical Practice Update on Diet and Nutritional Therapies in Patients With Inflammatory Bowel Disease: Expert Review. Gastroenterology. 2024 Mar;166(3):521–32.

4. Bischoff SC, Escher J, Hébuterne X, Kłęk S, Krznaric Z, Schneider S, et al. ESPEN practical guideline: Clinical Nutrition in inflammatory bowel disease. Clinical Nutrition. 2020 Mar;39(3):632–53.

5. Richman E, Rhodes JM. Review article: evidence-based dietary advice for patients with inflammatory bowel disease. Aliment Pharmacol Ther. 2013 Nov;38(10):1156–71.

6. Lee D, Albenberg L, Compher C, Baldassano R, Piccoli D, Lewis JD, et al. Diet in the Pathogenesis and Treatment of Inflammatory Bowel Diseases. Gastroenterology. 2015 May;148(6):1087–106.

7. Whelan K, Murrells T, Morgan M, Cummings F, Stansfield C, Todd A, et al. Food-related quality of life is impaired in inflammatory bowel disease and associated with reduced intake of key nutrients. The American Journal of Clinical Nutrition. 2021 Apr;113(4):832–44.

8. Lewis JD, Abreu MT. Diet as a Trigger or Therapy for Inflammatory Bowel Diseases. Gastroenterology. 2017 Jan;152(2):398–414.e6.

9. Limdi JK, Aggarwal D, McLaughlin JT. Dietary Practices and Beliefs in Patients with Inflammatory Bowel Disease: Inflammatory Bowel Diseases. 2016 Jan;22(1):164–70.

10. Tinsley A, Ehrlich OG, Hwang C, Issokson K, Zapala S, Weaver A, et al. Knowledge, Attitudes, and Beliefs Regarding the Role of Nutrition in IBD Among Patients and Providers: Inflammatory Bowel Diseases. 2016 Oct;22(10):2474–81.

11. Bernstein KI, Promislow S, Carr R, Rawsthorne P, Walker JR, Bernstein CN. Information needs and preferences of recently diagnosed patients with inflammatory bowel disease: Inflammatory Bowel Diseases. 2011 Feb;17(2):590–8.

12. Limdi JK, Butcher RO. Information resources and inflammatory bowel disease: Inflammatory Bowel Diseases. 2011 Aug;17(8):E89–90.

13. Cima RR, Anderson KJ, Larson DW, Dozois EJ, Hassan I, Sandborn WJ, et al. Internet use by patients in an inflammatory bowel disease specialty clinic: Inflammatory Bowel Diseases. 2007 Oct;13(10):1266–70.

14. Prince AC, Moosa A, Lomer MCE, Reidlinger DP, Whelan K. Variable access to quality nutrition information regarding inflammatory bowel disease: a survey of patients and health professionals and objective examination of written information. Health Expect. 2015 Dec;18(6):2501–12.

15. Yeo YH, Samaan JS, Ng WH, Ting PS, Trivedi H, Vipani A, et al. Assessing the performance of ChatGPT in answering questions regarding cirrhosis and hepatocellular carcinoma. Clin Mol Hepatol [Internet]. 2023 Mar 22 [cited 2023 Apr 5]; Available from: http://www.e-cmh.org/journal/view.php?doi=10.3350/cmh.2023.0089

16. Samaan JS, Yeo YH, Rajeev N, Hawley L, Abel S, Ng WH, et al. Assessing the Accuracy of Responses by the Language Model ChatGPT to Questions Regarding Bariatric Surgery. OBES SURG [Internet]. 2023 Apr 27 [cited 2023 May 29]; Available from: https://link.springer.com/10.1007/s11695-023-06603-5

17. Lechien JR, Georgescu BM, Hans S, Chiesa-Estomba CM. ChatGPT performance in laryngology and head and neck surgery: a clinical case-series. Eur Arch Otorhinolaryngol [Internet]. 2023 Oct 24 [cited 2023 Oct 28]; Available from: https://link.springer.com/10.1007/s00405-023-08282-5

18. Kaarre J, Feldt R, Keeling LE, Dadoo S, Zsidai B, Hughes JD, et al. Exploring the potential of ChatGPT as a supplementary tool for providing orthopaedic information. Knee Surg Sports Traumatol Arthrosc. 2023 Nov;31(11):5190–8.

19. Samaan JS, Margolis S, Srinivasan N, Srinivasan A, Yeo YH, Anand R, et al. Multimodal Large Language Model Passes Specialty Board Examination and Surpasses Human Test-Taker Scores: A Comparative Analysis Examining the Stepwise Impact of Model Prompting Strategies on Performance [Internet]. 2024 [cited 2024 Sep 21]. Available from: http://medrxiv.org/lookup/doi/10.1101/2024.07.27.24310809

20. Klang E, Sourosh A, Nadkarni GN, Sharif K, Lahat A. Evaluating the role of ChatGPT in gastroenterology: a comprehensive systematic review of applications, benefits, and limitations. Therap Adv Gastroenterol. 2023 Jan;16:17562848231218618.

21. Gravina AG, Pellegrino R, Cipullo M, Palladino G, Imperio G, Ventura A, et al. May ChatGPT be a tool producing medical information for common inflammatory bowel disease patients’ questions? An evidence-controlled analysis. World J Gastroenterol. 2024 Jan 7;30(1):17–33.

22. Sciberras M, Farrugia Y, Gordon H, Furfaro F, Allocca M, Torres J, et al. Accuracy of Information given by ChatGPT for Patients with Inflammatory Bowel Disease in Relation to ECCO Guidelines. Journal of Crohn’s and Colitis. 2024 Mar 23;jjae040.

23. Ghersin I, Weisshof R, Koifman E, Bar-Yoseph H, Ben Hur D, Maza I, et al. Comparative evaluation of a language model and human specialists in the application of European guidelines for the management of inflammatory bowel diseases and malignancies. Endoscopy. 2024 Sep;56(09):706–9.

24. Gong EJ, Bang CS. Evaluating the role of large language models in inflammatory bowel disease patient information. World J Gastroenterol. 2024 Aug 7;30(29):3538–40.

25. Naqvi HA, Delungahawatta T, Atarere JO, Bandaru SK, Barrow JB, Mattar MC. Evaluation of online chat-based artificial intelligence responses about inflammatory bowel disease and diet. European Journal of Gastroenterology & Hepatology. 2024 Sep;36(9):1109–12.

26. openai. ChatGPT: Optimizing Language Models for Dialogue. 2023; https://openai.com/blog/chatgpt/. Accessed 1/1/2023, 2023.

27. OpenAI. GPT-4 Technical Report. 2023 [cited 2023 Aug 11]; Available from: https://arxiv.org/abs/2303.08774

28. Ouyang L, Wu J, Jiang X, Almeida D, Wainwright CL, Mishkin P, et al. Training language models to follow instructions with human feedback. 2022 [cited 2023 Feb 10]; Available from: https://arxiv.org/abs/2203.02155

29. King RC, Samaan JS, Yeo YH, Peng Y, Kunkel DC, Habib AA, et al. A Multidisciplinary Assessment of ChatGPT’s Knowledge of Amyloidosis: Observational Study. JMIR Cardio. 2024 Apr 19;8:e53421.

30. King RC, Samaan JS, Yeo YH, Mody B, Lombardo DM, Ghashghaei R. Appropriateness of ChatGPT in Answering Heart Failure Related Questions. Heart, Lung and Circulation. 2024 Sep;33(9):1314–8.

31. Samaan JS, Yeo YH, Ng WH, Ting PS, Trivedi H, Vipani A, et al. ChatGPT’s ability to comprehend and answer cirrhosis related questions in Arabic. Arab Journal of Gastroenterology. 2023 Sep; S1687197923000588.

32. Cunningham E. Are Low-Residue Diets Still Applicable? Journal of the Academy of Nutrition and Dietetics. 2012 Jun;112(6):960.

33. Larussa T, Suraci E, Marasco R, Imeneo M, Abenavoli L, Luzza F. Self-Prescribed Dietary Restrictions are Common in Inflammatory Bowel Disease Patients and Are Associated with Low Bone Mineralization. Medicina. 2019 Aug 20;55(8):507.

34. Marsh A, Kinneally J, Robertson T, Lord A, Young A, Radford-Smith G. Food avoidance in outpatients with Inflammatory Bowel Disease – Who, what and why. Clinical Nutrition ESPEN. 2019 Jun;31:10–6.

35. Godala M, Gaszyńska E, Durko Ł, Małecka-Wojciesko E. Dietary Behaviors and Beliefs in Patients with Inflammatory Bowel Disease. JCM. 2023 May 14;12(10):3455.

36. Habashi P, Bouchard S, Nguyen GC. Transforming Access to Specialist Care for Inflammatory Bowel Disease: The PACE Telemedicine Program. Journal of the Canadian Association of Gastroenterology. 2019 Oct 9;2(4):186–94.

37. Borren NZ, Conway G, Tan W, Andrews E, Garber JJ, Yajnik V, et al. Distance to Specialist Care and Disease Outcomes in Inflammatory Bowel Disease: Inflammatory Bowel Diseases. 2017 Jul;23(7):1234–9.

38. Malter L, Jain A, Cohen BL, Gaidos JKJ, Axisa L, Butterfield L, et al. Identifying IBD Providers’ Knowledge Gaps Using a Prospective Web-based Survey. Inflammatory Bowel Diseases. 2020 Aug 20;26(9):1445–50.

39. Bickmore TW, Pfeifer LM, Paasche-Orlow MK. Using computer agents to explain medical documents to patients with low health literacy. Patient Education and Counseling. 2009 Jun;75(3):315–20.

40. Liu JJ, Abraham BP, Adamson P, Barnes EL, Brister KA, Damas OM, et al. The Current State of Care for Black and Hispanic Inflammatory Bowel Disease Patients. Inflammatory Bowel Diseases. 2023 Feb 1;29(2):297–307.

41. Al Shamsi H, Almutairi AG, Al Mashrafi S, Al Kalbani T. Implications of Language Barriers for Healthcare: A Systematic Review. Oman Med J. 2020 Mar;35(2):e122.

42. Divi C, Koss RG, Schmaltz SP, Loeb JM. Language proficiency and adverse events in US hospitals: a pilot study. Int J Qual Health Care. 2007 Apr;19(2):60–7.

43. Yeo YH, Samaan JS, Ng WH, Ma X, Ting PS, Kwak MS, et al. GPT-4 outperforms ChatGPT in answering non-English questions related to cirrhosis [Internet]. Gastroenterology; 2023 May [cited 2023 Jul 29]. Available from: http://medrxiv.org/lookup/doi/10.1101/2023.05.04.23289482

44. Zhu Z, Ying Y, Zhu J, Wu H. ChatGPT’s potential role in non-English-speaking outpatient clinic settings. DIGITAL HEALTH. 2023 Jan;9: 20552076231184091.

